# Periodontopathogens in oral cancer: a meta-analysis of bacterial taxa of the oral microbiome associated with risk factors of oral squamous cell carcinoma

**DOI:** 10.1101/2022.03.11.22272244

**Authors:** Abdus Salam, Faisal Khan

**Affiliations:** CECOS-RMI Precision Medicine Lab, RMI Laboratory Building, Hayatabad Phase 5, Hayatabad, Peshawar 25000, Pakistan; Institute of Integrative Biosciences, CECOS University, Phase 6, Hayatabad, Peshawar, Pakistan; Department of Bioengineering, University of Engineering and Applied Science, Swat, Pakistan

**Keywords:** Oral squamous cell carcinoma, Microbial dysbiosis, Risk factors, Periodontitis

## Abstract

This study aims to investigate the distribution of microbial taxa that are present in abundance in the oral cavity of patients diagnosed with Oral Squamous Cell Carcinoma (OSCC). We begin with a search for relevant literature on the OSCC microbiome in electronic databases (PubMed and Google Scholar). From the identified literature, studies were considered for data extraction based on an inclusion criteria according to PRISMA guidelines. From an initial 1217 published studies, a total of 15 relevant studies were identified that fulfilled the inclusion criteria. These studies were conducted for the detection of microbial taxa in the oral cavities of patients with OSCC by correlation with healthy controls for differential microbial abundance. The data from the selected studies provided evidence on microbial taxa in different anatomical sites of the oral cavity i.e. gingival region, tongue, buccal site and floor of the mouth. The most common method for the detection of microbial flora in the literature was 16s rRNA sequencing. Only those studies from the literature were considered for further analysis that showed the association of risk factors i.e. tobacco smoking and smokeless, betel quid, alcohol and periodontitis with OSCC. Risk factors in the resulting 6 studies showed a strong odd’s ratio (OR) with statistical significance (p-value <0.05). The calculated risk ratio (RR) of these risk factors also demonstrated substantial heterogeneity. These studies showed an increase in the abundance of periodontopathogens belonging to the genus *Fusobacterium, Capnocytophaga, Prevotella, Parvimonas* and *Porphyromonas*. The microbial taxa associated in abundance with risk factors of OSCC such as smoked or smokeless tobacco, betel quid and alcohol were quite similar to the microbial taxa that cause periodontitis. The detection for abundance of periodontopathogens in OSCC a class of putative biomarkers at early stages of tumor development in OSCC, in individuals exposed to these risk factors.

## Introduction

Oral Squamous Cell Carcinoma (OSCC) commonly known as oral cancer is a subset of Head and Neck Squamous Cell Carcinoma (HNSCC) that has been classified as “lips, oral cavity and pharynx” by the International Classification of Disease (https://icd.who.int). OSCC accounts for an estimated 354,864 new cases resulting in 177,384 deaths annually and has less than 50% 5-year survival rate in most regions of the world ^1^. The major risk factors associated with OSCC are the use of tobacco (both smoke and smokeless), betel quid, alcohol consumption, periodontitis and Human Papilloma Virus (HPV) infection ^2,3^. However, the number of OSCC cases is growing in which the above-mentioned risk factors may not yet be linked but rather suggested to be associated with microbial dysbiosis ^4-6^. Microbial dysbiosis is a term that describes the imbalance in the composition of microbial communities or its relative abundance that resides in or on the host’s body and has an influence on the patient’s health. Not only has microbial dysbiosis been reported to be associated with cancer^(11-13)^ but also with obesity ^7^, diabetes ^8^, cardiovascular diseases^9^ and inflammatory bowel diseases ^10^.

The hypothesized role played by microbial dysbiosis in cancer is chronic inflammation ^11^, activation of T-cells ^12^, secretion of oncoproteins and toxic metabolites ^13^ which potentially leads to DNA damage. Recently, interest in microbial dysbiosis in the oral cavity has increased with the aim of uncovering its role in OSCC progression and development. A significant amount of research on OSCC microbiome has been carried out in the last couple of decades using a wide range of experimental techniques for the identification of microbial population in both cases and controls as well as in different control types, i.e. matched-pair and healthy controls ^14^. These studies have led to the recognition of different microbial taxa that are enriched in OSCC samples. However, the detected microbial taxa in these studies do not overlap neatly possibly due to the differences in methodology, sample sources or control types and also because of exposure to different risk factors, cultural differences, geography, diet and cancer stages ^15,16^. But, on the other hand, different microbial tax in a given environmental niche have been shown to be performing a conserved function or exhibiting functional redundancy^17,18^.

The key microbial species that have been identified most frequently in OSCC cases belong to genus *Fusobacterium, Prevotella, Porphyromonas, Capnocytophaga* and *Peptostreptococcus*^19^. Several studies have already been conducted to explore the biological function of these taxa in OSCC progression ^5,20^. These enriched biological functions include chronic inflammation induced by either lipopolysaccharide, interleukin 1 beta, interleukin-8, toll-like receptors, genomic repair system, bacterial mobility, chemotaxis and flagellar assembly, cytoskeleton, and carbohydrate-related metabolism ^15,16,21-24^. Due to these biological functions, the imbalance in microbial taxa or microbial dysbiosis in the oral cavity has been linked to initiating and progressing OSCC.

Risk factors such as tobacco smoking, smokeless tobacco, betel quid, alcohol drinking and periodontitis have been previously shown to have an association with microbial dysbiosis ^14,16,21^. Some studies have also shown that these risk factors are associated with the enrichment of microbial taxa that cause periodontitis. The study by Grine *et al* ^25^ and Jiang *et al* ^26^ showed that the relative abundance of periodontopathogens elevates in smoking as compared to the non-smoking group. The study by Halboub *et al* revealed that participants consuming smokeless tobacco (shammah) showed an increase in abundance of microbial genera that causes periodontitis as well as the synthesis of acetaldehydes that might be involved in oral carcinogenesis ^27^. It has been also shown by Fan *et al* ^28^ and Valles *et al* ^29^ that both smoking and smokeless tobacco as well as alcohol consumption might be performing a similar role in altering the microbial diversity in the oral cavity that might lead to periodontitis and oral cancer. The link between periodontitis and oral cancer has also been explored by the transcriptomics study undertaken by Deng *et al* which showed the transcriptional activation of genes by periodontopathogens that promote carcinogenesis and might eventually lead to oral cancer ^30^.

The enrichment of microbial taxa in OSCC patients that are exposed to the risk factors have been reported in several studies. However, there has been no systematic effort to develop a comprehensive picture of microbial taxa in OSCC cases associated with these risk factors. In this review, an online database (PubMed) was explored for the identification of studies relevant to risk factors associating with microbial dysbiosis in OSCC specifically bacteriome and retrieve patient-level data of exposure to these risk factors. The aim of this study is to identify bacterial taxa that have been reported to increase in abundance in OSCC patients exposed to tobacco (both smoking and smokeless), alcohol and periodontitis and also to assess the strength of each association. These bacterial signatures might be beneficial for further study as diagnostic biomarkers for oral squamous cell carcinoma cases associated with these risk factors.

## Materials and Methods

### Design for Literature Selection

We used the Preferred Reporting Items for Systematic Reviews and Meta-Analysis (PRISMA) ^31^ guidelines as a benchmark for the selection of literature and for data extraction (Figure 1). The main objective of this study was to single out bacterial taxa that were identified by differential abundance in studies where OSCC patients were evidently exposed to risk factors. To select relevant literature, terms such as *Oral Squamous Cell Carcinoma, Microbiome, Bacteriome, Microbiota, Microbial Diversity, Microbial Dysbiosis* and *Metagenomics* were taken together with boolean operators such as “AND” and “OR”. Table 1 lays out the list of queries that was used for filtering in PubMed and Google Scholar (as of July 8th, 2020). The searching criteria was further narrowed down by confining organisms to *Homo sapiens* and language to English. In addition, manual searches were performed to identify supplementary studies from systematic studies published in Al-Hebshi *et al* ^14^, Gopinath *et al* ^32^ and Robledo-Sierra *et al* ^33^.

**Table 1.**
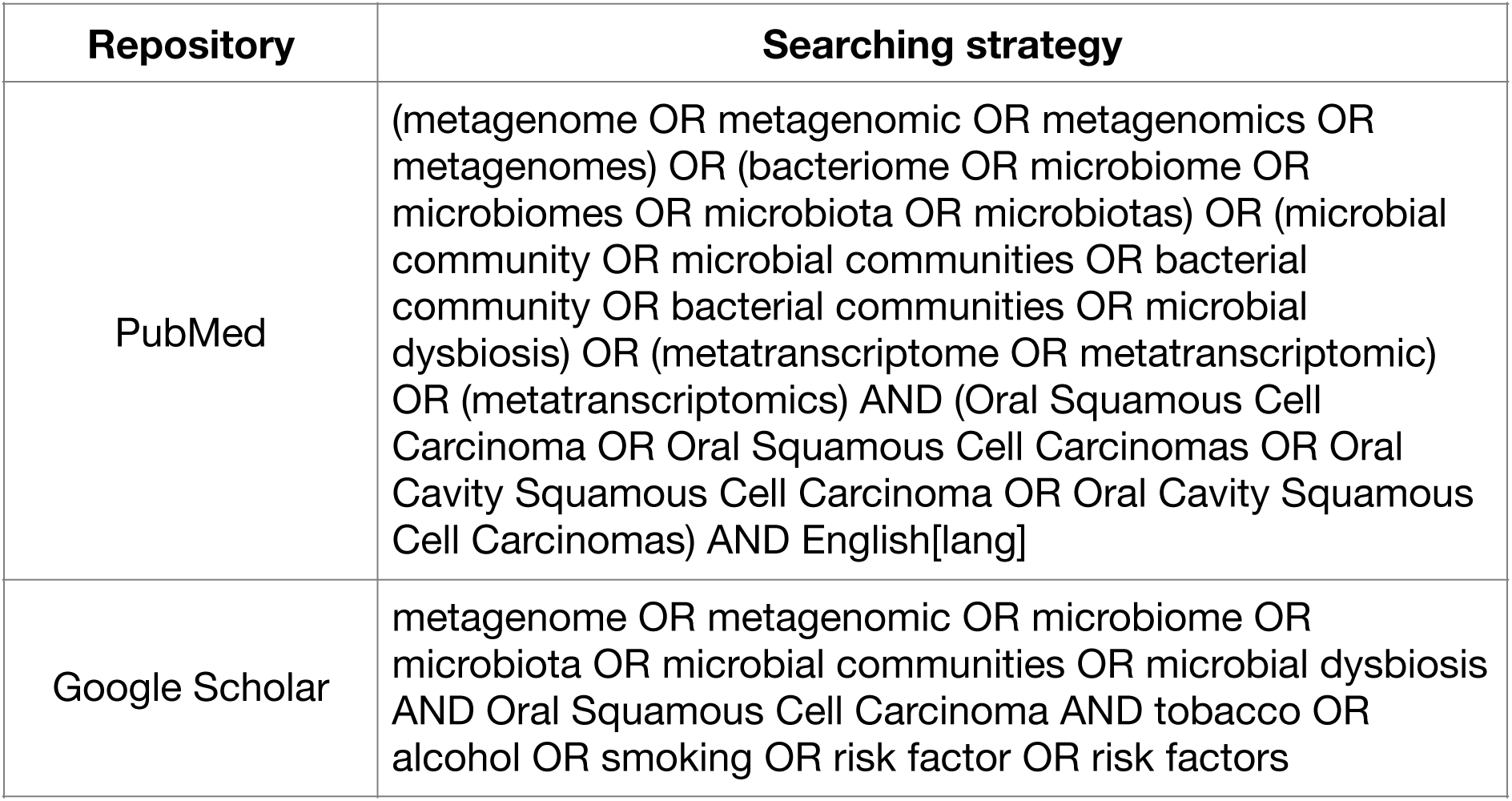
Searching strategy for relevant published studies.

**Figure 1.**
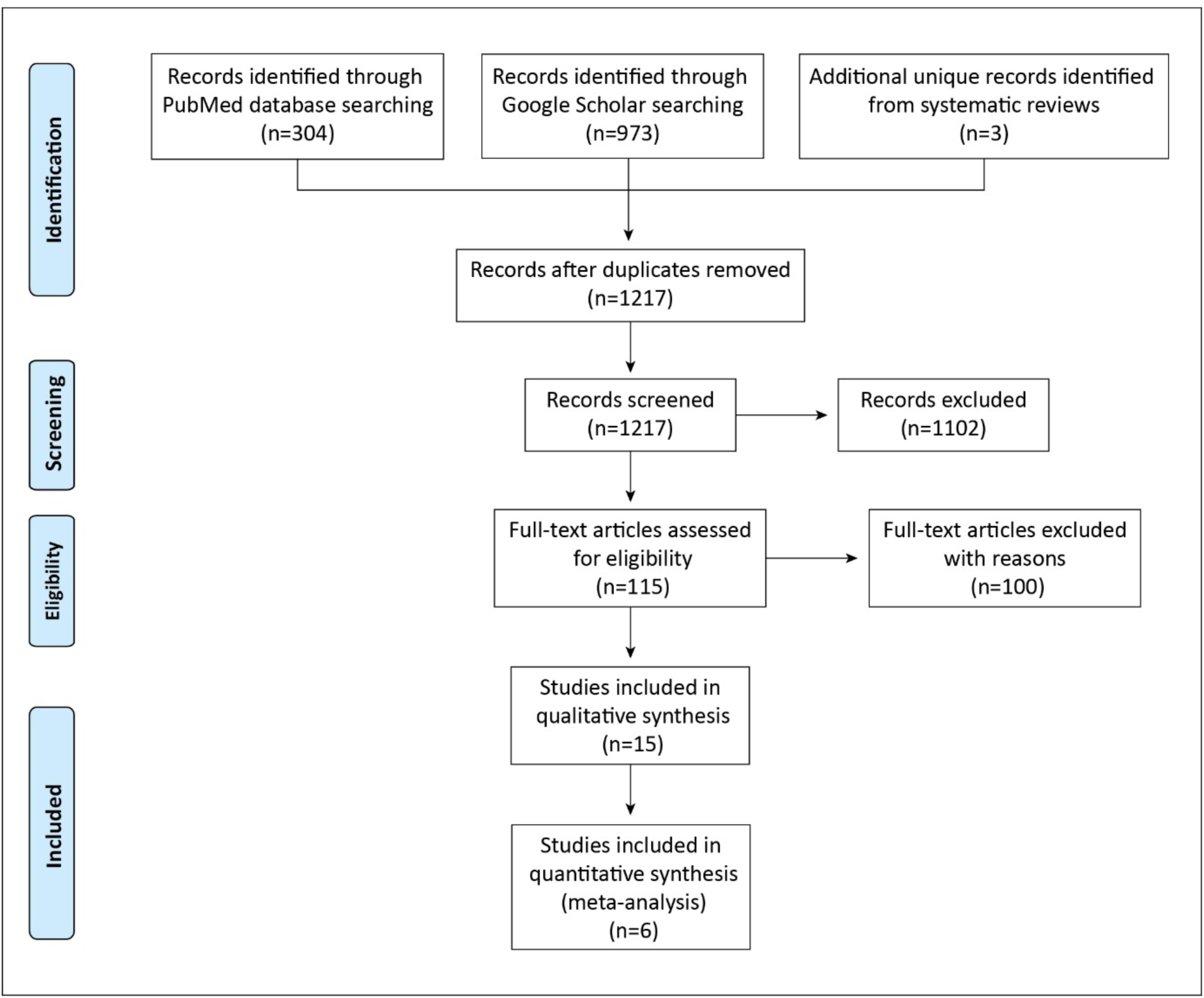
A flow diagram of systematic analysis in accordance with the PRISMA guidelines. A search for literature has been performed using three main sources; (a) PubMed, (b) Google Scholar, and (c) Review articles. Furthermore, the identified literature has been filtered by removing duplicates and including studies of interest for quantitative analysis.

### Inclusion Criteria for Selected Studies

After removing duplicates found in each search, the titles and abstracts of unique articles were initially screened for microbial taxa detection in OSCC. In the case where information in the title and abstract was insufficient, the full-text of the articles were reviewed prior to being either included or excluded. The remaining articles were included only if, (1) the samples were extracted from OSCC and not from squamous cell carcinomas of other anatomic sites of the head and neck region, (2) the studies reported a differential abundance of microbial taxa by correlating cases with controls, (3) the control sample was extracted from healthy participants and (4) the detected microbial taxa were bacteria. Studies not meeting the above conditions were excluded.

### Data Characterization of the Selected Studies

Studies meeting the inclusion criteria were gathered and thoroughly reviewed. A summary of key attributes such as name of the first author, publication date, country of origin, methodology used, tumor sites, sample size of cases and controls, participants’ mean age, sample types of cases and controls, and the number of participants exposed to risk factors are presented in Table 2. Additionally, bacterial taxa that have been reported by differential abundance in cases and controls are characterised in Table S1.

**Table 2.**
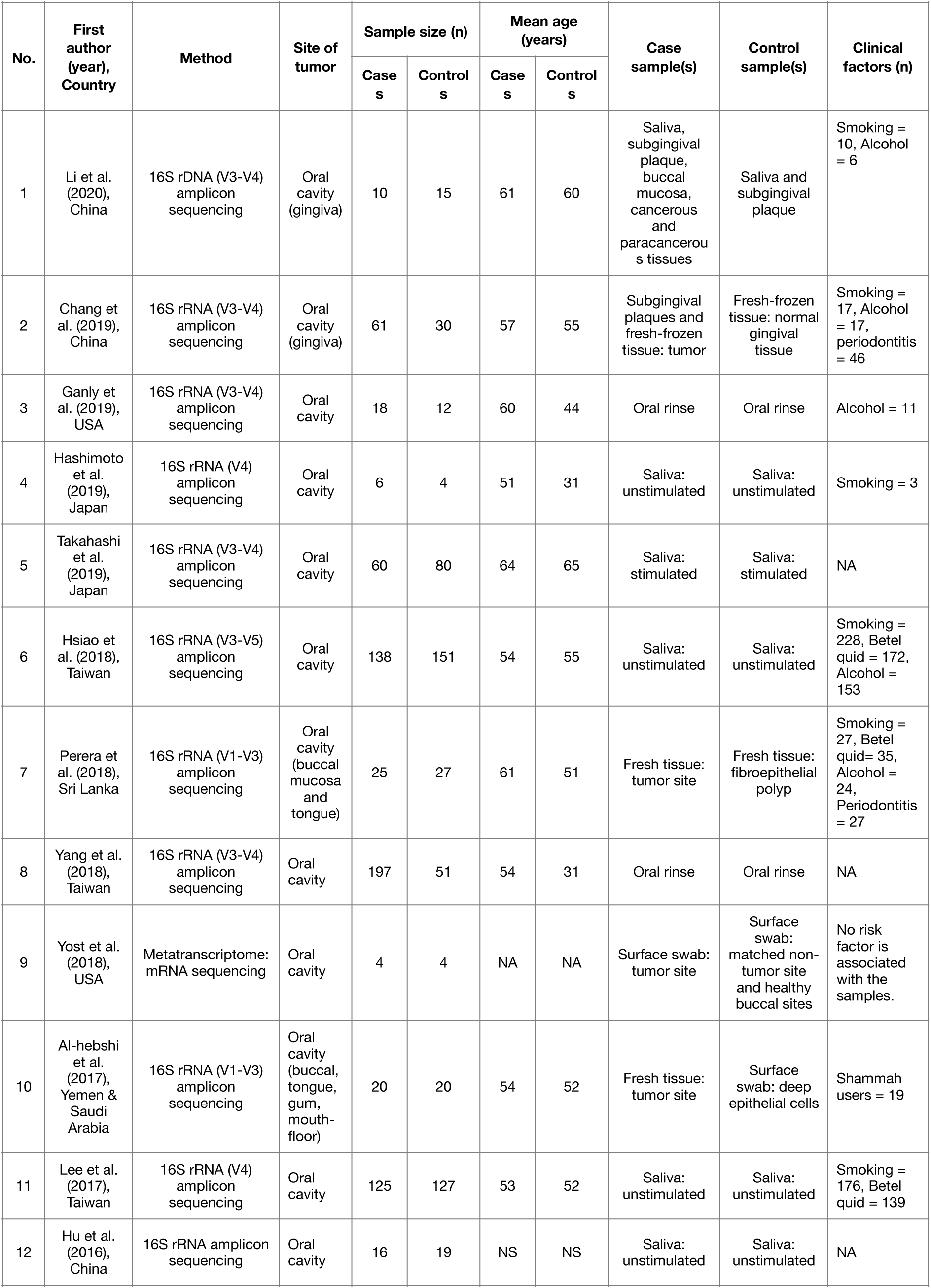
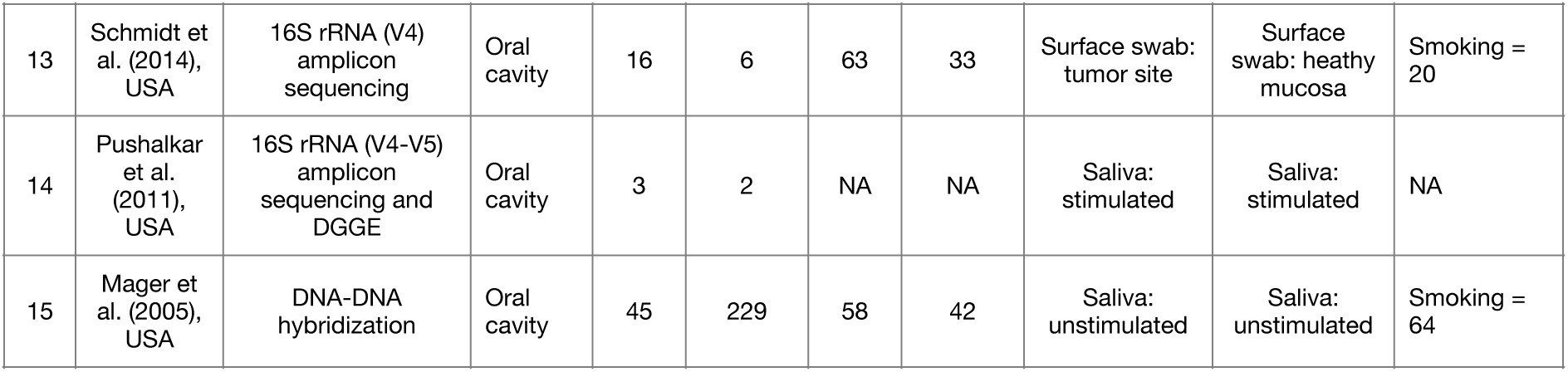
General characteristics of the included studies.

### Statistical Analysis

For quantitative analysis, only those studies that have clearly mentioned the number of participants in case and control groups that were exposed to risk factors were considered. The odds ratio (OR) for each study was calculated using MedCalc (https://www.medcalc.org/calc/). The statistics are shown in Table 3. Studies having OR greater than 1 and p-value less than 0.05 were considered for estimating risk ratio (RR). Figure 2 depicts a forest plot of the RR analysis that was performed using “Metafor” (package of R v3.6)^34^.

**Table 3.**
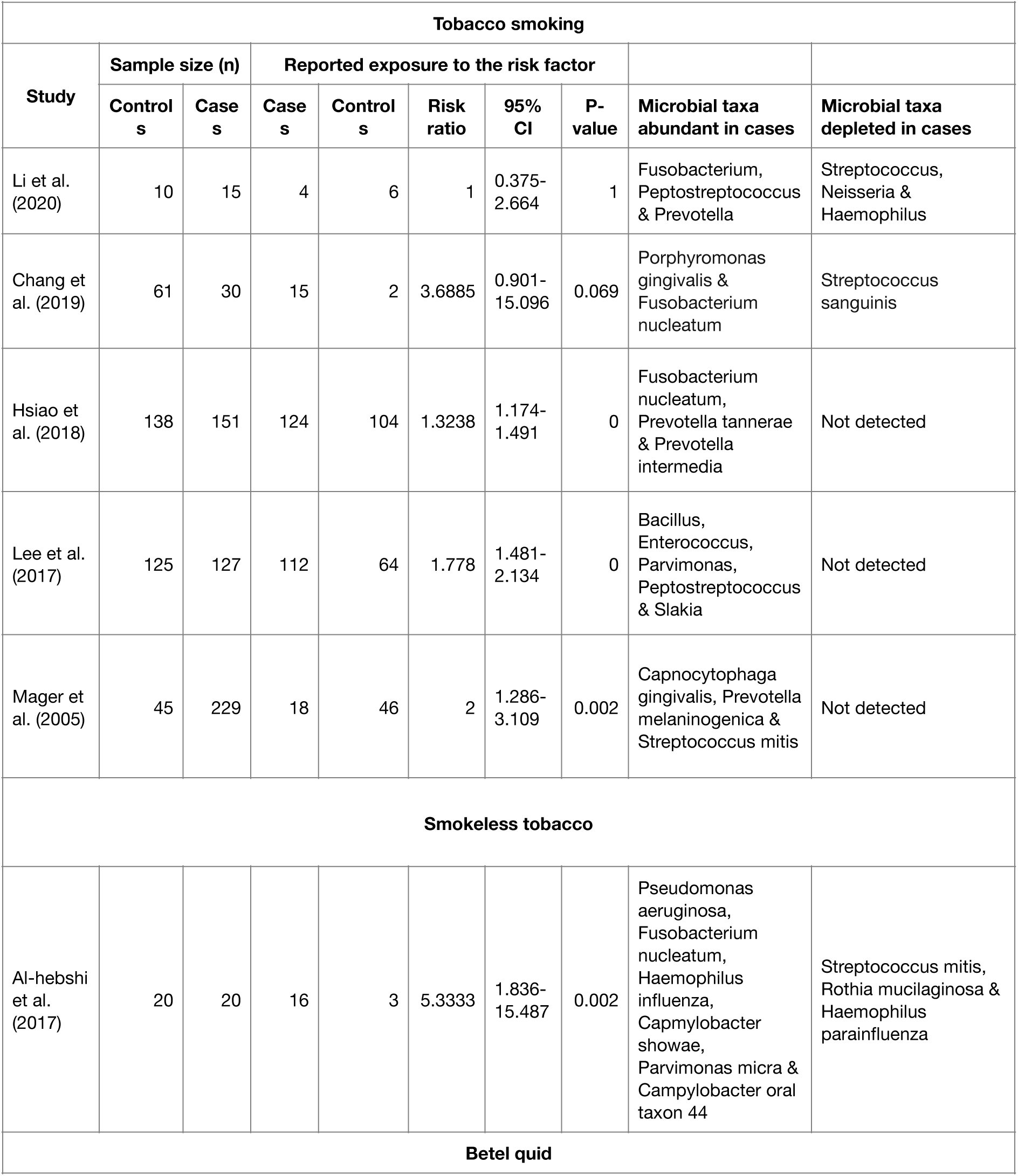
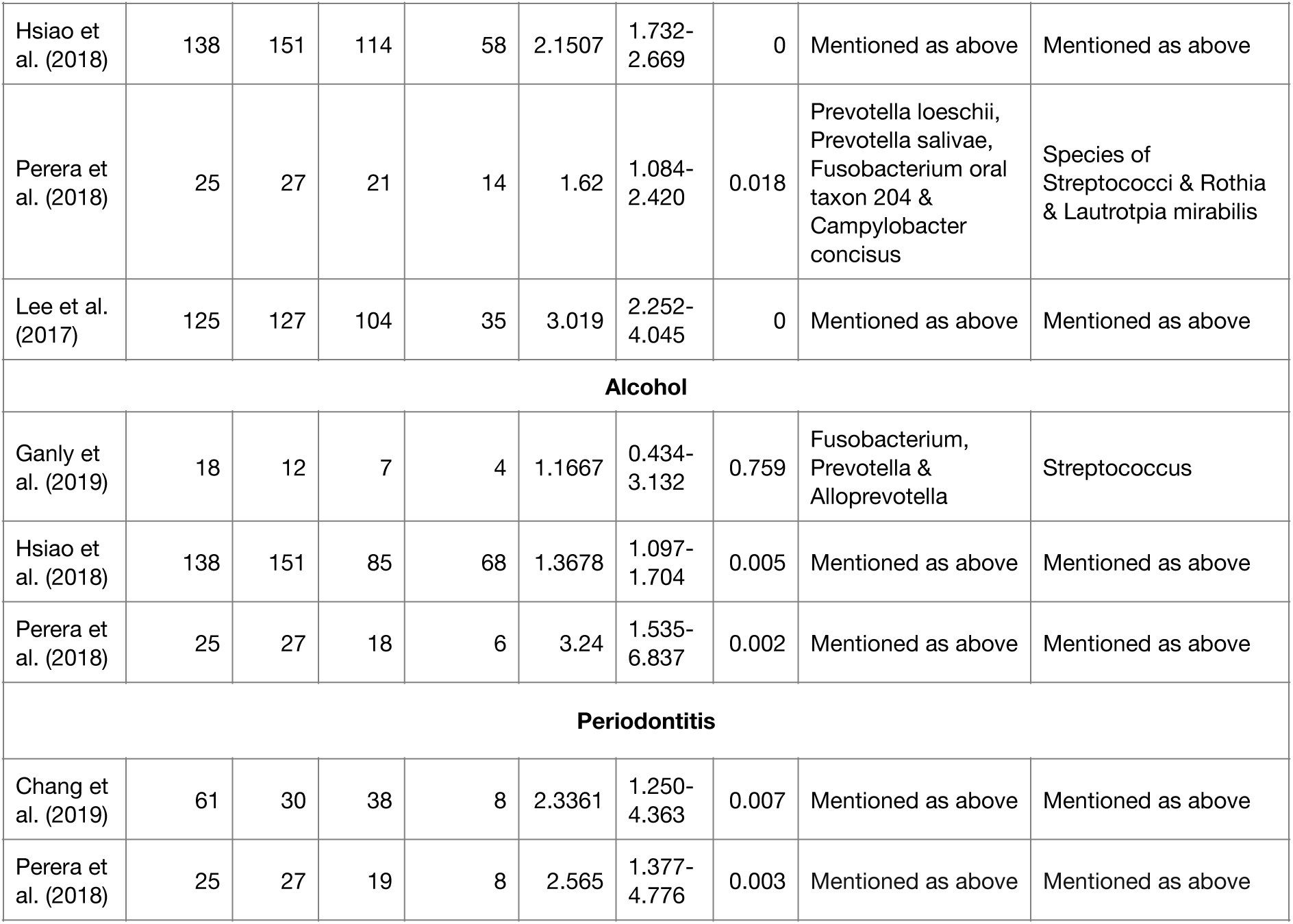
Selection of studies based on its strength of association with the risk factors.

**Figure 2.**
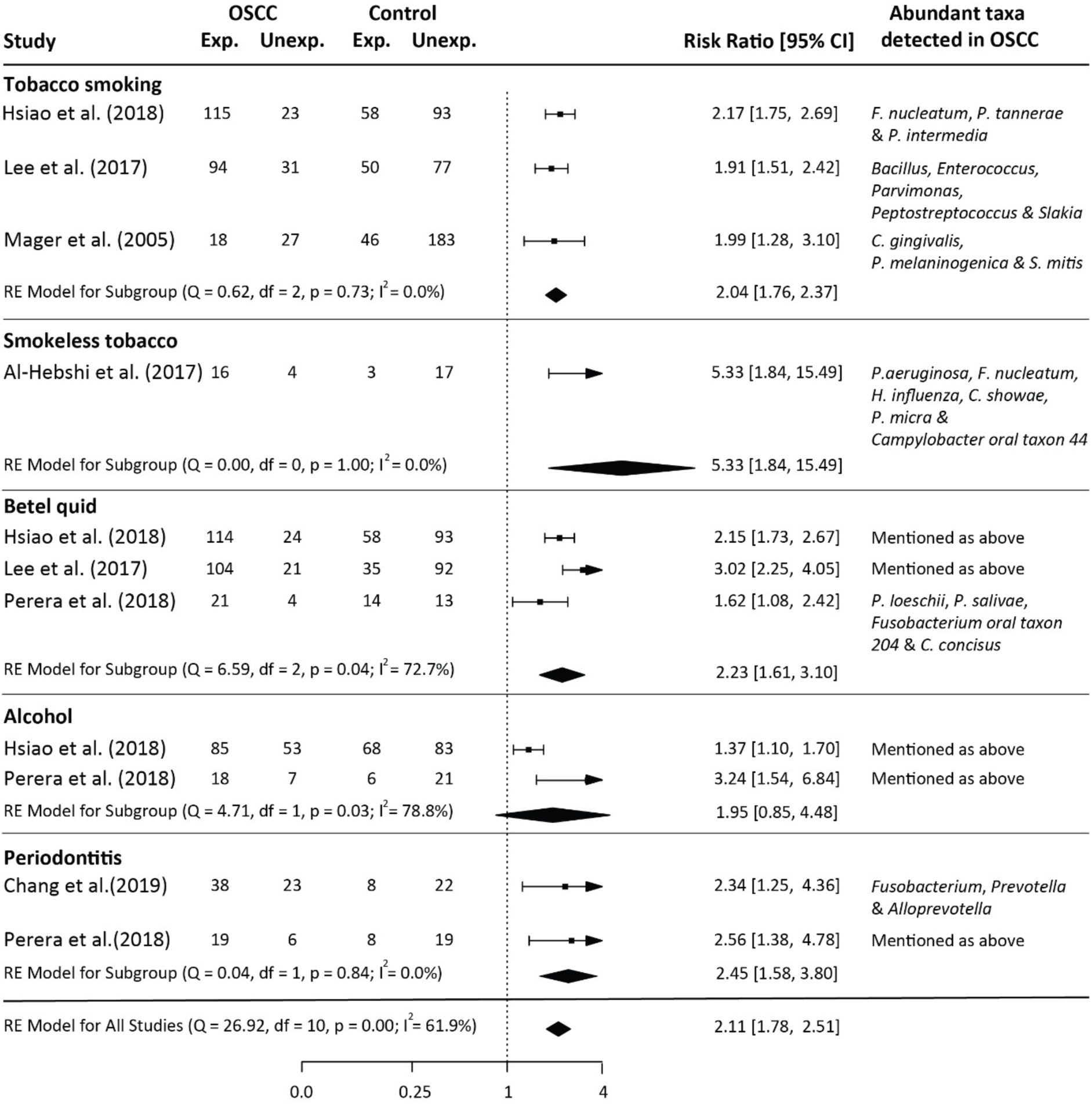
Quantitative analysis of the selected studies using risk ratio. The quantitative analysis has been evaluated for each risk factor category; Tobacco smoking, Smokeless tobacco, Betel quid, Alcohol and Periodontitis as well as combined.

## Results

### Characterisation of Relevant Studies

A total of 1214 unique articles were found after a preliminary search looking through PubMed and Google Scholar. Additionally, three more studies by Al-Hebshi *et al* ^14^, Gopinath *et al* ^35^ and Robledo-Sierra *et al* ^33^ were also considered, making it a total of 1217 articles. A total of 1102 were excluded after perusal due to irrelevance to the theme of the study. Furthermore, a total of 100 studies were excluded after reading their abstracts. Only 15 articles met the inclusion criteria and were considered for qualitative analysis. PRISMA guidelines were followed for strategising the selection and design of the study (Figure 1). These studies identified the relative abundance of microbial taxa by comparing OSCC cases with healthy controls. Table 1 provides a summary of key details of the final selection of studies.

The selected studies were published between 2005 to 2020 and the sampling population were primarily from Americas and Asia. The samples were extracted from participants of diverse ethnicity and gender except for the study of Perera *et al* ^21^, Hsiao *et al* ^16^, Yost *et al* ^24^, Al-Hebshi *et al* ^22^, and Pushalkar *et al* ^36^, where both cases and controls were extracted from males. The mean age of the participants across all studies with OSCC and controls was 57.5 years and 45.6 years respectively; whereas, the mean age of the participants with OSCC and healthy controls in studies by Hashimoto *et al* ^37^, Ganly *et al* ^38^, Yang *et al* ^15^ and Schmidt *et al* ^39^ showed mean age difference of more than 15 years. Thirteen out of all fifteen studies used 16s rRNA amplicon sequencing for the detection of microbial abundance while the remaining two used DNA-DNA hybridization^40^ and meta-transcriptomics^24^ approaches, respectively. The V3-V4 region was the most frequently used for microbial taxa identification in the case of 16s amplicon sequencing approach.

The sample size of OSCC participants and controls across all studies ranged from 3 to 197 and 2 to 229, respectively (Table 2). Samples were extracted from the oral cavity of both tumor and controls including the anatomic sites of gingival region, tongue, buccal site, and floor of the mouth. Saliva (stimulated and unstimulated), gingival plaque, buccal mucosa, fresh tissues, and surface swabs were used for sampling from OSCC cases; whereas, the types of samples extracted from controls included saliva (stimulated and unstimulated), fresh tissues, and buccal mucosa. Only Li *et al* ^41^, Ganly *et al* ^38^, Chang *et al* ^42^, Lee *et al* ^43^ and Schmidt *et al* ^39^ analysed differential microbial abundance in more than two groups which also included pre-malignant lesions and periodontal cohorts.

### Discovery of frequent Periodontopathogens in OSCC samples

Microbial taxa reported in the included studies correlating OSCC with controls were retrieved from individual studies at either species or genus taxonomic level as shown in Table S1. A total of 23 bacterial genera that were detected by differential abundance were reported in the included studies. The reported genera were categorized into five phyla:

*Bacteroidetes, Firmicutes, Fusobacteria, Proteobacteria* and *Actinobacteria*, of which the first three phyla were in abundance. *Fusobacteria*, which is the most frequently detected phylum of bacteria, has been reported by several studies in OSCC ^21,22,41^ to have increased in abundance. *Firmicutes* and *Bacteroidetes*, on the other hand, were found abundant across different studies in both cases and controls.

Studies conducted by Hsiao *et al* ^16^, Yost *et al* ^24^, Lee *et al* ^43^, Pushalkar *et al* ^36^ and Mager *et al* ^40^ did not report abundance of bacterial taxa in controls. At genus level, *Streptococcus* is the most frequently reported microbial taxa to have increased in abundance in both cases and controls. *Fusobacterium, Prevotella, Parvimonas* and *Capnocytophaga* were found abundantly in OSCC while *Rothia* and *Neisseria* were depleted. Furthermore, other genera such as *Porphyromonas, Gemella, Peptostreptococcus, Haemophilus, Filifibacter, Micrococcus, Lactobacillus, Leptotrichia, Abiotrophia, Veillonella*, and *Campylobacter* were found in a small number of studies that showed abundance in either OSCC or controls or both.

At specie level, *Fusobacterium nucleatum* and *Poprphromonas gingivalis* were reported in two studies while *Fusobacterium periodonticum, Prevotella tannerae* and *Prevotella intermedia, Prevotella melaninogenica, Prevotella loeschii, Prevotella salivae, Campylobacter concisus* and *Campylobacter showae, Peptostreptococcus anaerobius, Pseudomonas aeruginosa, Filfibacter alocis, Capnocytophaga gingivalis, Parvimonas micra* and *Haemophilus influenzae* were reported to have an increase in abundance in cases, in one study each. However, the abundance of *Streptococcus mitis* has been detected in abundance in cases by Mager *et al* ^40^ while the study of Al-Hebshi *et al* ^22^ showed its abundance in controls. Furthermore, the abundance *Streptococcus sanguinis, Lautropia mirabilis, Rothia mucilaginosa* and *Haemophilus parainfluenzae* were reported to deplete in cases only.

### Risk Factors Associated with OSCC and Changes in Microbial Abundance: A Quantitative Analysis

Studies that reported risk factor exposure for each participant in both cases and controls, were considered for further quantitative analysis. Takahashi *et al* ^44^, Yang *et al* ^15^, Yost *et al* ^24^, Hu *et al* ^45^ and Pushalkar *et al* ^36^ were excluded due to the insufficient information of participants exposure to risk factors. Similarly, Hashimoto *et al* ^37^ (n=10) and Schmidt *et al* ^39^ (n=22) were excluded due to small sample size (>25). Participants in both cases and controls, in the remaining eight studies, conducted by Li *et al* ^41^, Chang *et al* ^42^, Hsiao *et al* ^16^, Perera *et al* ^21^ and Lee *et al* ^43^ indicated that both groups were exposed to more than one risk factor. Whereas, Ganly *et al* ^38^, Al-Hebshi *et al* ^22^ and Schmidt *et al* ^39^, each exhibited the exposure of participants to a single but different risk factor.

From the eight studies that were selected for quantitative analysis, the number of participants exposed to risk factors in both cases and controls were retrieved. Participants that were either currently exposed to risk factors or were so in the past were considered exposed. Whereas, those participants which were reported by the authors of the selected studies as occasionally exposed were considered unexposed. In two studies, Chang *et al* ^42^ and Perera *et al* ^21^, periodontitis was considered as a risk factor in participants having severe or moderate periodontitis disease. The Odds Ratio was calculated to estimate the strength of association between risk factors in the selected eight studies as shown in Table 3. Studies were categorized in five groups: tobacco smoking, smokeless tobacco, betel quid, alcohol and periodontitis, based on the differences in types of risk factors that were associated with participants. In Li *et al* ^41^ and Chang *et al* ^42^, where participants were exposed to tobacco smoking, and Ganly *et al* ^38^, where participants were exposed to alcohol, were not statistically significant (p-value > 0.05). These studies were removed prior to further analysis. Studies by Hsiao *et al* ^16^, Lee *et al* ^43^ and Mager *et al* ^40^, having participants exposed to tobacco smoking, Al-Hebshi *et al* ^22^, having participants exposed to smokeless tobacco, Hsiao *et al* ^16^, Perera *et al* ^21^ and Lee *et al* ^43^, having participants exposed to alcohol consumption, and Chang *et al* ^42^ and Perera *et al* ^21^, having exposure to periodontitis displayed OR value > 1 and p-value less than 0.05.

Figure 2 depicts a forest plot of the RR analysis that was performed for the five studies that displayed significant OR and p-value. The analyses considered using the random-effect (RE) model for each risk factor individually as well as for all the reported risk factors collectively. The collective analysis showed a substantial heterogeneity measure (*I*^2^) of almost 62% with p-value < 0.00 and RR of 2.11. Al-Hebshi *et al* ^22^, associating its participants with smokeless tobacco (shammah, showed a higher RR value of 5.33 and lower heterogeneity (I^2^ = 0.0%) while the studies of Hsiao *et al* ^16^ and Perera *et al* ^21^, where the participant are alcohol-consuming, showed lower RR value of 1.95 and higher heterogeneity measure (I^2^ = 78.8%). Furthermore, Hsiao *et al* ^16^, Lee *et al* ^43^ and Mager *et al* ^40^, having tobacco smoking participants, and Hsiao *et al* ^16^, Lee *et al* ^43^ and Perera *et al* ^21^, having betel quid consuming participants, showed RR value of 2.04 and lower homogeneity measure (I^2^ = 0.0%), and 2.45 and higher homogeneity (I^2^ = 72.7%), respectively.

The studies conducted by Al-Hebshi *et al* ^22^, Hsiao *et al* ^16^, Lee *et al* ^43^ and Mager *et al* ^40^ where participants were exposed to all risk factors except periodontitis, showed an increase in bacterial abundance of periodontopathogens belonging to the genus *Fusobacterium, Prevotella, Porphyromonas, Capnocytophaga* and *Parvimonas*. At the species level, periodontopathogens that were found in to be in enriched in exposed participants were *F. nucleatum, P. gingivalis, C. gingivalis, P. micra* and several other species of *Prevotella*. These bacterial taxa have also been reported to have an association with chronic inflammation by triggering the expression of proinflammatory molecules and later causing periodontitis ^11,46^. Furthermore, they were also reported in Chang *et al* ^42^ and Perera *et al* ^21^ where participants were exposed to periodontitis as a risk factor.

## Discussions

Commensal microbial communities in human gut and oral cavity perform a wide range of biological functions that are associated with nutrition, immune modulation, and metabolism ^47-49^. Changes in their diversity have been observed and reported to have a role in carcinogenesis as well as progression of cancer ^50-52^. It has also been suggested that the risk factors of cancer demonstrate an interplay with the microbial diversity during carcinogenesis ^53,54^. In this study, we evaluated microbial taxa that reportedly increased in abundance in previously published studies relating to risk factors associating with OSCC. The studies were retrieved from online databases and screened based on an inclusion criteria.

Microbial diversity is a commonly used index that considers the presence of variable or distinguished phylotypes. Microbial diversity in the 15 studies we selected do not overlap strongly. This might be due to the ethnicity and age of participants, nature of the sample and different anatomic sites in the oral cavity from which the sample were collected^32^. From the selected studies, information relating to microbial taxa was retrieved which belonged to five different phyla: *Bacteroidetes, Firmicutes, Fusobacteria, Proteobacteria* and *Actinobacteria*. Species belonging to the Phyla *Firmicutes, Proteobacteria*, and *Bacteroidetes* were found with variations in both cases and controls. However, the members of phylum *Fusobacteria* showed an increase in abundance when compared to the members of other phyla. Interestingly, the study of Yang *et al* ^15^ showed that with the progression of cancer’s stage, the abundance of *Fusobacteria* significantly increased while the abundance level of *Actinobacteria* and *Bacteroidetes* diminished. The members of *Actinobacteria* have been also shown to inhibit tumorigenesis on the mucosal layer and its depletion might be playing a vital role in promoting carcinogenesis ^55,56^.On the basis of frequency, microbial genera that were found to increase in abundance were *Fusobacterium, Prevotella, Parvimonas, Capnocytophaga* and *Porphyromonas. Fusobacterium* is the most frequently detected bacterial genus in OSCC and an increase in its abundance has been systematically evaluated by Bronzato *et al* ^57^ in OSCC studies. Yang *et al* ^15^ demonstrated that at the different stages of cancer, with the progression of OSCC, the abundance of *Fusobacterium* increases while *Streptococcus, Haemophilus, Actinomyces* and *Porphyromonas* decreases. *Fusobacterium* has been shown to be involved in the coaggregation of microbial species, helping in survival and colonization of pathogenic microbes in the oral cavity and hence could be responsible for metastasizing lesions ^58-61^. *F. nucleatum* was the most reported bacterial species while others such as *F. noviforme* and *F. Periodonticum* were also reported to increase in abundance in OSCC ^15^. Other than OSCC, the increase in abundance of *F. nucleatum* has been also demonstrated in gastrointestinal ^62-64^ and colorectal cancer ^62,65^. In colorectal cancer, *F. nucleatum* has been assumed to have a role in cellular proliferation pathways, invasion of colorectal cells’ environment and suppression of the host’s immune system ^66,67^.

High level members from genus *Prevotella* and *Parvimonas* have been most commonly seen in colorectal cancer ^68,69^ as well as in OSCC but its role in promoting carcinogenesis still remains unreported. *C. gingivalis* and *P. gingivalis* are periodontopathogens and its enrichment has been frequently observed in OSCC ^21,42^. The hypothesized role of *C. gingivalis* and *P. gingivalis* is the activation of the inflammatory cascade pathway that leads to the cause of chronic periodontitis and might be involved in initiating OSCC ^70^. Genus *Rothia* and *Neisseria* showed depletion in OSCC while the abundance in *Streptococcus* species remained inconsistent across different cases as it increased in some cases and depleted in others. Furthermore, the depletion of *Rothia* and *Neisseria* has also been seen in breast cancer survivors and esophageal cancer but their actual role still needs to be identified ^71,72^.

In the included studies, information related to the risk factors i.e. tobacco smoking, smokeless tobacco, betel quid, alcohol and periodontitis were retrieved. HPV is a risk factor associated with microbial dysbiosis in oropharyngeal squamous cell carcinoma ^73-75^ however, it was not found in the included studies. Besides that, we could not find a single study that could report the association of microbial taxa in OSCC with participants that are not exposed to any of the risk factors. The quantitative analysis of the included studies showed a significant association between OSCC patients and their exposure to the risk factors. It has been also reported that persistent exposure to these risk factors results in microbial dysbiosis that ultimately increases the risk of oral epithelial dysplasia and eventually leads to OSCC ^76^. The bacterial taxa associated with the risk factors, except periodontitis, that were reported to increase in abundance belonged to genus *Fusobacterium, Prevotella, Campylobacter, Capnocytophaga, Porphyromonas* and *Parvimonas* in OSCC showed similarity to participants with periodontitis.

It has been previously reported that tobacco is a confounding factor having association with both periodontitis and oral cancer ^77^. However, in the included studies it was also found that the risk factors such as tobacco (both smoking and smokeless), betel quid and alcohol might be confounding factors for both periodontitis and OSCC. Hsiao *et al* ^16^ has also shown the link between an increase in the abundance of periodontopathogenic bacteria and OSCC risk. It has also been suggested that these periodontopathogens can be used as biomarkers not only for periodontitis and OSCC but also for detection of cardiovascular ^78,79^, autoimmune diseases ^80^ and several other types of cancers ^81,82^. Nonetheless, further studies, that include a larger size of participants from different ethnicities, better taxonomic resolution as well as the exposure of participants to a particular rather than the collective risk factors in each group, are still needed.

## Conclusions

Imbalances in microbial abundance or dysbiosis of bacteriome in the oral cavity have been previously reported to have a link with OSCC. However, shifting in microbial abundance in association with risk factors such as tobacco smoking, smokeless tobacco, betel quid, alcohol and periodontitis in OSCC has not been explored yet. These risk factors might be involved in the development of an optimal environment for pathogenic bacteria that causes inflammation and periodontitis and leading to the proliferation of OSCC. In this study, the microbial dysbiosis that are associated with risk factors of OSCC shows an increase in the abundance of periodontopathogens and inflammatory bacteriome such as *Fusobacterium, Capnocytophaga, Prevotella, Parvimonas* and *Porphyromonas* and the depletion of *Rothia* and *Neisseria*. It was also found that consumable risk factors of OSCC i.e. tobacco, betel quid and alcohol are involved in increasing the risk of periodontitis and could be a confounding with OSCC. It means that the development of periodontitis in participants exposed to these risk factors could also be a significant indicator of the initiation and progression of OSCC. However, further studies are needed to explore the role of risk factors in the development of OSCC. Also, the bacterial taxa could be used as biomarkers for the detection of OSCC at early stages.

## Data Availability

All data produced in the present work are contained in the manuscript.

## Author Contributions

Abdus Salam contributed to the conception, design, and drafted the manuscript; Faisal Khan contributed to conception and critically revised the manuscript. Both authors gave final approval and agreed to be accountable for all aspects of the work ensuring integrity and accuracy.

## Acknowledgements

The authors declare no potential conflicts of interest with respect to the authorship and/or publication of this article. We are very grateful to Saad Zaheer for reviewing and helping in rewriting the manuscript.

## Conflict of interest

None

---

**Supplementary table 1.** Bacterial taxonomy that are reported as enriched or depleted in the selected studies.

| Study | Detected taxonomy level | Abundant in OSCC cases | Depleted in OSCC cases |
| --- | --- | --- | --- |
| Li et al. (2020) | Genus | Fusobacterium, Peptostreptococcus and Prevotella | Streptococcus, Neisseria and Haemophilus |
| Chang et al. (2019) | Specie | Fusobacterium nucleatum and Porphyromonas gingivalis | Streptococcus sanguinis |
| Ganly et al. (2019) | Genus | Fusobacterium, Prevotella and Alloprevotella | Streptococcus |
| Hashimoto et al. (2019) | Genus | Prevotella, Haemophilus and Solobacterium | Rothia, Streptococcus, Leptotrichia, Gemella and Neisseria |
| Takahashi et al. (2019) | Genus | Fusobacterium, Capnocytophaga, Alloprevotella and Peptostreptococcus | Rothia and Haemophilus |
| Hsiao et al. (2018) | Specie | Fusobacterium nucleatum, Prevotella tannerae and Prevotella intermedia | Not detected |
| Perera et al. (2018) | Genus/Specie | Prevotella loeschii, Prevotella salivae, Fusobacterium oral taxon 204 and Campylobacter concisus | Species of Streptococci, Rothia and Lautropia mirabilis |
| Yang et al. (2018) | Genus/Specie | Fusobacterium, Parvimonas, Streptococcus, Haemophilus, Filifibacter, Fusobacterium periodonticum, Parvimonas micra, Streptococcus constellatus, Haemophilus influenzae and Filifibacter alocis. | Streptococcus, Haemophilus, Porphyromonas and Veillonella |
| Yost et al. (2018) | Genus/Specie | Species of Fusobacteria and Streptococci, Prevotella, Porphyromonas gingivalis and Peptostreptococcus anaerobius | Not detected |
| Al-Hebshi et al. (2017) | Specie | Fusobacterium nucleatum, Pseudomonas aeruginosa, Haemophilus influenza, Campylobacter showae, Parvimonas micra and Campylobacter oral taxon 44 | Streptococcus mitis, Rothia mucilaginosa and Haemophilus parainfluenza |
| Lee et al. (2017) | Genus | Bacillus, Enterococcus, , Parvimonas, Peptostreptococcus and Slakia | Not detected |
| Hu et al. (2016) | Genus | Bacillus | Streptococcus and Abiotrophia |
| Schmidt et al. (2014) | Genus | Fusobacterium and Prevotella | Streptococcus and Rothia |
| Pushalkar et al. (2011) | Genus | Streptococcus, Rothia, Gemella, Peptostreptococcus, Lactobacillus, Micrococcus and Porphyromonas | Not detected |
| Mager et al. (2005) | Specie | Capnocytophaga gingivalis, Prevotella melaninogenica and Streptococcus mitis | Not detected |

## Notes

### Competing Interest Statement

The authors have declared no competing interest.

### Funding Statement

The study was undertaken by members of the Precision Medicine Lab at the National Centre for Big Data and Cloud Computing funded by the Higher Education Commission, Pakistan.

